# Incident autoimmune diseases in association with a SARS-CoV-2 infection: A matched cohort study

**DOI:** 10.1101/2023.01.25.23285014

**Authors:** Falko Tesch, Franz Ehm, Annika Vivirito, Danny Wende, Manuel Batram, Friedrich Loser, Simone Menzer, Josephine Jacob, Martin Roessler, Martin Seifert, Barbara Kind, Christina König, Claudia Schulte, Tilo Buschmann, Dagmar Hertle, Pedro Ballesteros, Stefan Baßler, Barbara Bertele, Thomas Bitterer, Cordula Riederer, Franziska Sobik, Lukas Reitzle, Christa Scheidt-Nave, Jochen Schmitt

## Abstract

**Objectives:** To investigate whether the risk of developing an incident autoimmune disease is increased in patients with previous COVID-19 disease compared to people without COVID-19.

**Method:** A cohort was selected from German routine health care data covering 38.9 million individuals. Based on documented diagnoses, we identified individuals with polymerase chain reaction (PCR)-confirmed COVID-19 through December 31, 2020. Patients were matched 1:3 to control patients without COVID-19. Both groups were followed up until June 30, 2021. We used the four quarters preceding the index date until the end of follow-up to analyze the onset of autoimmune diseases during the post-acute period. Incidence rates (IR) per 1000 person-years were calculated for each outcome and patient group. Poisson models were deployed to estimate the incidence rate ratios (IRRs) of developing an autoimmune disease conditional on a preceding diagnosis of COVID-19.

**Results:** In total, 641,704 patients with COVID-19 were included. Comparing the incidence rates in the COVID-19 (IR=15.05, 95% CI: 14.69-15.42) and matched control groups (IR=10.55, 95% CI: 10.25-10.86), we found a 42.63% higher likelihood of acquiring autoimmunity for patients who had suffered from COVID-19. This estimate was similar for common autoimmune diseases, such as Hashimoto thyroiditis, rheumatoid arthritis, or Sjögren syndrome. The highest IRR was observed for autoimmune disease of the vasculitis group. Patients with a more severe course of COVID-19 were at a greater risk for incident autoimmune diseases.

**Conclusions:** SARS-CoV-2 infection is associated with an increased risk of developing new-onset autoimmune diseases after the acute phase of infection.

## Introduction

To date, numerous research articles have been published on COVID-19, the acute disease that results from infection with severe acute respiratory syndrome coronavirus 2 (SARS-CoV2) and its chronic counterpart long/post-COVID. SARS-CoV-2 is a positive-polar single-stranded RNA (ssRNA) virus that is transmitted directly by respiratory droplets[1]. The primary target of the virus is the lung epithelium. From there, it can enter the peripheral blood and infect cells of tissues expressing the ACE2 protein, such as the heart, kidneys, gastrointestinal tract and brain, causing a multiorgan manifestation[1]. First, SARS-CoV-2 induces a type I and type III interferon-based antiviral response at the cellular level and robust production of chemokines that recruit inflammatory cells, e.g., monocytes and neutrophils[2]. An acute SARS-CoV-2 infection is characterized by an increase in proliferating, metabolically hyperactive plasmablasts (PBs) producing first IgM antibodies and later IgG antibodies[2]. An early occurrence of SARS-CoV-2-specific cytotoxic CD8+ T cells is correlated with effective viral clearance and mild disease[3].

After the acute phase of infection, some people may develop long-lasting symptoms, known as post-COVID. This is defined by signs and symptoms that develop during or after SARS-CoV-2 infection that are consistent with COVID-19, last more than 12 weeks and cannot be explained by an alternative diagnosis[4]. Most studies thus far have focused on symptoms that partly wane over time [5-9]. Many studies examined a small selective sample of patients, and only a few studies included a control group or information on chronic health conditions, such as SARS-CoV-2 infection[10]. This evidence is insufficient to estimate the burden of COVID-19 in a population and the resulting future challenges to healthcare systems. Routinely collected health data, such as claims data from health insurances, offer large sample sizes to investigate rare but potentially severe outcomes in this context.

To date, different respiratory, cardiovascular, neurological and mental diseases as well as various symptoms in the context of long/post-COVID have been studied with routine health care data [11-18]. The group of autoimmune diseases is less discussed in the literature, although autoantibodies could be found in patients after SARS-CoV-2 infection, e.g., anti-type I IFNs, anti-IFN-α and anti-nuclear antibodies (ANAs) [19]. So far there is limited evidence on newly manifested autoimmune diseases after an infection based on several case reports and one recent cohort study using UK health record data [20-22]. Furthermore, COVID-19 itself shares some similarities with systemic autoimmune rheumatic diseases, which could be a challenge for diagnostics [23, 24].

Here, we investigated the association of COVID-19 and a set of incident autoimmune diseases in a large cohort of patients enrolled in German statutory health insurances.

## Methods

### Study design

We conducted a matched cohort study based on routine health care data as applied in a previous study [16]. In the present analysis, we compared the rates of newly diagnosed autoimmune disease between individuals with and without documented SARS-CoV-2 infection. Persons infected with SARS-CoV-2 during 2020 and matched controls were followed until June 30, 2021, for a minimum of three and a maximum of 15 months using the date of COVID-19 onset as the index date for randomly selected match groups. Following the NICE guidelines on long COVID [25] and the clinical case definition of post-COVID-19 conditions proposed by the World Health Organization (WHO)[4], we defined the post-COVID-19 phase starting three months after the initial diagnosis of COVID-19. Outpatient services are documented per quarter rather than on a daily basis in the German statutory health care billing system. We therefore considered a diagnosis to have been made in the post-COVID-19 phase if it was newly documented in the second quarter after the index date or later. This operationalization ensures a time interval of at least three months between the date of COVID-19 diagnosis and post-COVID-19 outcome incidence.

### Cohorts

The COVID-19 cohort included individuals with polymerase chain reaction (PCR)-confirmed COVID-19 diagnosis (ICD-10 U07.1) in 2020. To calculate risk exposure time, we defined the onset of COVID-19 inside the quarter as the index date by using the date of an outpatient PCR test or the date of admission to a hospital with a COVID-19 diagnosis. In rare cases where no PCR test had been billed to the insurance company and no hospital stay was recorded, other documented events, such as the start of sick leave or the first contact with the responsible physician, served to determine the index date. The control cohort included individuals who were not diagnosed with ICD-10 U07.1 or ICD-10 U07.2 without a documented COVID-19 diagnosis between January 1st, 2020, and June 30th, 2021.

We excluded individuals with COVID-19 diagnosis without laboratory virus detection (ICD-10-GM: U07.2) from the COVID-19 groups and non-COVID-19 controls to reduce distortions due to misclassification. We further excluded individuals who were not continuously registered with the respective health insurance company between 2019-01-01 (or birth) and 2021-06-30 (or death), whichever came first, because relevant outcomes and preexisting health conditions may not be visible in our data. For each individual, preexisting medical conditions could be assessed for at least 12 months prior to the matching point of the COVID-19 and control cohorts. Starting from the index date, which was assigned from the COVID-19 case, matched individuals were jointly followed for a maximum of 15 months. This permitted comparison of two groups over the same period to compare their risk of developing any of the predefined incident autoimmune diseases conditional on COVID-19.

### Data

The underlying data sources were set up for the “Post-COVID-19 Monitoring in Routine Health Insurance Data” (POINTED) consortium [16] to study the long-lasting effects of the COVID-19 pandemic in Germany. The POINTED consortium is coordinated by the Center for Evidence-Based Healthcare (ZEGV) at the TU Dresden and consists of the German National Public Health Authority, the Robert Koch Institute, health research institutes and statutory health insurances. It is funded partly by the German Federal Ministry of Health (BMG).

We used routine health care data from different German statutory health insurances: Techniker Krankenkasse, BARMER, DAK Gesundheit, IKK classic, AOK PLUS, and several company health insurance funds (InGef[26]). In total, these data cover approximately 39 million individuals, which corresponds to nearly half of the total German population. In addition to sociodemographic characteristics (age and sex) and vital status (via the date of death), we had access to comprehensive information on healthcare utilization in the outpatient and inpatient health care sectors. The data comprise records on diagnoses (according to the International Statistical Classification of Diseases and Related Health Problems - German Modification, ICD-10-GM), medical procedures (according to the “Operationen-und Prozedurenschluessel”, OPS; German modification of the International Classification of Procedures in Medicine, ICPM), information on outpatient medical services (according to “Einheitlicher Bewertungsmassstab”, EBM), and prescribed medications (according to the German Anatomical Therapeutic Chemical (ATC) Classification). Changes in the medical condition of an individual were determined subject to the quarterly information available in the electronic health care data. Only patients from 2020 were selected, as this ensured that the effect was not influenced by vaccinations.

### Matching

To minimize differences between the COVID-19 and control cohorts in terms of covariates that may confound relationships between outcomes and exposure, we applied 1:3 matching with replacement for COVID-19 to non-COVID-19 patients. For each individual in the COVID-19 cohort, we selected three non-COVID-19 individuals with identical age (in years), sex and whether or not an autoimmune disease was present before the index date. We chose exact matching on these characteristics to facilitate stratified analysis. In addition, we accounted for the presence of covariates by propensity score matching. The estimation of the propensity score was based on logistic regression including all insured individuals. Given different sets of prevalent medical conditions considered as covariates, we estimated separate regression models for children/adolescents and adults.

After matching individuals with COVID-19 and controls, we excluded individuals from the match groups if they died before the beginning of the post-COVID phase, i.e., within the quarter of the COVID-19 diagnosis or the following quarter. We also excluded individuals with COVID-19 who lacked a matching partner. When analyzing specific health outcomes, we further excluded individuals from the analysis if the considered outcome was documented in two of the four quarters preceding in the outpatient setting or once in the inpatient setting. To maintain balance of cohorts regarding covariates, we excluded a complete match group of COVID-19 and control cases if the outcome was preexisting for the individual with COVID-19 or all of his or her matched non-COVID-19 control cases. For estimation, we weighted data from individuals in the control cohort with the inverse number of individuals remaining in the respective match group (i.e., weights between 1/3 and 1) to ensure that total weights in the control cohort added up to the number of individuals in the COVID-19 cohort.

### Health outcomes

Based on the clinical experience of the author team, we defined 64 potential outcomes from 41 autoimmune diseases studied during follow-up 3 to 15 months after documented COVID-19 infection, e.g., assigned index date. Operationalization of these outcomes was based on inpatient and outpatient diagnoses according to ICD-10-GM and the guidelines good practice secondary data analysis (GPS) of the German Society for Epidemiology (DGEpi)[27]. In 23 cases, a more specific definition of the outcome with suitable medication was chosen. For type I diabetes, only those cases with an insulin prescription were considered valid. A complete list of the considered outcomes and their definitions can be found in supplementary material S1.

### Covariates

We used information on preexisting chronic conditions as available from 2019 health records to adjust for potential confounders in the relationship of exposure (COVID-19) and incident autoimmune diseases. The approach is similar to a previous study [16].

For each individual, we used information on preexisting health conditions in the four quarters preceding the index date. The 13 prevalent morbidities for children/adolescents and 34 prevalent morbidities were based on published evidence and clinical expertise (supplementary material S2). In addition, we included age, sex and the number of recorded inpatient and outpatient contacts as covariates. In line with previous studies[11], we included the severity of COVID-19 as a stratification feature and differentiated between 1) individuals with outpatient diagnoses of COVID-19, 2) individuals with a hospital visit with COVID-19, and 3) individuals with intensive care and/or mechanical ventilation with COVID-19.

### Statistical analyses

The incidence rates (IRs) of autoimmune diseases per 1000 person-years were estimated. Differences in IRs between COVID-19 and non-COVID-19 patients were estimated using Poisson regression models to estimate incidence rate ratios (IRRs). As a prerequisite, we derived aggregated information on each health outcome by counting incident cases of the respective autoimmune disease within the COVID-19 and control groups. Since the number of incident cases for each outcome varied across the match groups, we assigned weights to the remaining control cases that added up to 1. The pooling of individual-level data was not possible due to data protection restrictions. The different insurance datasets were therefore analyzed separately by authorized institutes or the health care research departments within the respective health insurances. Each authorized institute calculated the required aggregate statistics and provided them to ZEGV, where regressions based on combined aggregate data were performed.

To synthesize evidence across datasets, we note that point estimates from aggregate matched data are equal compared to the case of Poisson regression based on individual level data[28]. The characteristics of Poisson regression applied to aggregate count data allowed for consistent estimation of incidence rates regardless of the distribution of the outcome on the individual level when the conditional mean function is correctly specified[29]. However, variance estimates from aggregates tend to be larger, meaning that the statistical significance of the presented effects may be underestimated. Utilizing a main advantage of Poisson regression, we adjusted for differences in individual-specific times at risk (time between the index date and the end of the observation period or death) due to inclusion of these times as offset in the model. Stratified aggregation enabled us to deploy separate estimators for age, sex, prevalent autoimmune disease, and severity of COVID-19. We conducted all analyses using the statistical programming language R [30].

## Results

### Description of the study population

In 2020, 38.9 million individuals were insured in one of the participating insurance companies for at least one day. We excluded persons not continuously enrolled in 2019 (n=2,074,654) or between January 1st, 2020 and June 30, 2021 (n=2,051,855), those with a COVID-19 diagnosis without definite laboratory confirmation (ICD-10 U07.2) (n=3,549,324), and those with a COVID-19 diagnosis in the first two quarters of 2021 (n=569,410) from the analyses (Figure 1). From the remaining sample, 670,301 individuals with a COVID-19 diagnosis were matched 1:3 to controls. For 29 individuals with COVID-19 (0.004%), no suitable matching partner was found. After matching, there were 28,810 individuals with COVID-19 and 20,932 control cases who died in the time between the (assigned) index date and the beginning of the second quarter following the (assigned) index quarter. By excluding these cases from the post-COVID-19 observable cohort, an additional 81,570 controls (from 28,810 match groups) and 55 individuals with COVID-19 were left without matching partners and thus had to be excluded from the study. The final study population consisted of 641,407 individuals with COVID-19 and 1,560,357 non-COVID-19 individuals serving as controls in 1,907,992 control cases. Most of the individuals in the COVID-19 group had three control cases. Only minor fractions of individuals with COVID-19 had two (n=13,187; 2.06%) or one (n=869; 0.14%) control case/s. As sampling with replacement was conducted, the control cohort consisted of only 1,560,357 distinct individuals (Figure 1).

**Figure 1.**
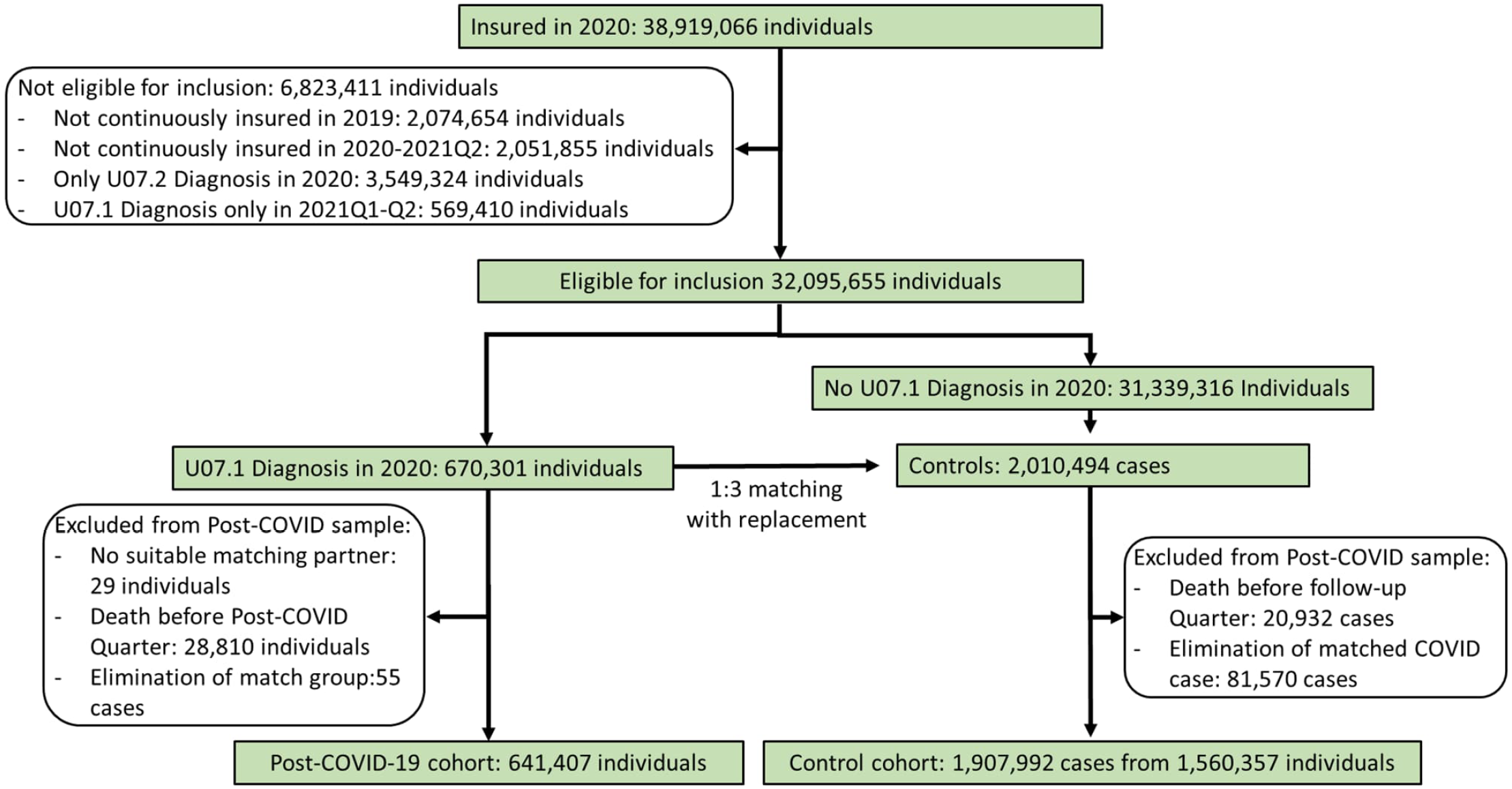
Flowchart for the selection of the COVID-19 and control groups

### Description of the population

Among the 641,407 COVID-19 individuals included, 76,518 (11.9%) had a prevalent autoimmune disease prior to COVID-19. Among those without any preexisting autoimmune diagnoses, 6,489 developed a first autoimmune disease 3 to 15 months after SARS-CoV-2 infection. Of the individuals with preexisting autoimmunity, 1744 developed an additional autoimmune disease. More than half of those in the COVID-19 and control cohorts were female (57.3%); the majority were between 18 and 64 years of age (74.2% and 74.6%), while 9.4% and 9.5% were below 18 years of age and 16.3% and 15.9% were 65 years of age or older, respectively. Regarding the course of the acute COVID-19 disease, 6.4% of individuals were hospitalized, and 1.6% received intensive care treatment and/or mechanical ventilation (Table 1).

**Table 1.** Characteristics of COVID-19 and control cohort after matching.

| Category | n COVID-19 | Percent COVID-19 | n Controls | Percent Control |
| --- | --- | --- | --- | --- |
| Total | 641,407 | 100 | 1,907,992 | 100 |
| Sex |  |  |  |  |
| Male | 273,868 | 42.7 | 1,092,873 | 42.7 |
| Female | 367,539 | 57.3 | 815,119 | 57.3 |
| Age |  |  |  |  |
| 0-17 | 60,535 | 9.4 | 181,210 | 9.5 |
| 18-64 | 476,085 | 74.2 | 1,423,132 | 74.6 |
| 65-79 | 60,889 | 9.5 | 180,060 | 9.4 |
| 80+ | 43,898 | 6.8 | 123,590 | 6.5 |
| Severity of COVID-19 |  |  |  |  |
| Outpatient | 590,204 | 92.0 | - | - |
| Hospital | 40,846 | 6.4 | - | - |
| ICU/Ventilation | 10,357 | 1.6 | - | - |
| Autoimmunity |  |  |  |  |
| Any preexisting autoimmune disease | 76,518 | 11.9 | 237,035 | 12.4 |
| First onset of autoimmune disease | 6,489 | 1.1 | 13376 | 0.8 |
| Additional autoimmune disease with preexisting autoimmunity | 1744 | 2.3 | 3324 | 1.4 |

Depending on the number of matched controls per individual with COVID-19, each entered the analysis with a weight between 1/3 and 1.

### Incidence of a new autoimmune disease

The incidence rate (IR) of any autoimmune disease 3 to 15 months after SARS-CoV-2 infection was 15.05 (95%-CI 14.69-15.42) per 1000 person-years in the COVID-19 group and 10.55 (95% CI: 10.25-10.86) in the control group for patients without a prior autoimmune disease. Hence, the excess risk due to SARS-CoV-2 infection was 4.50 per 1000 person-years. The IRR for an incident autoimmune disease was 1.43 (95%-CI=1.37-1.48). Furthermore, incident autoimmune diseases were more frequent among individuals with COVID-19 who had any preexisting autoimmune disease, while the relative likelihood to controls with preexisting autoimmune disease was smaller (IR COVID-19=38.10, IRR=1.23 95%-CI=1.15-1.32) (Table 2).

**Table 2.** Incidence rate (IR) and incidence rate ratio (IRR) for an autoimmune disease among persons with compared to those without SARS-CoV2 infection 3 to 15 months after the index date

| Outcomes | Events<br>COVID-19 | IR<br>COVID-19 | Events<br>Weighted<br>Controls* | IR<br>Weighted<br>Controls | IRR | 95%CI |
| --- | --- | --- | --- | --- | --- | --- |
| First onset of autoimmune disease | 6489 | 15.05 | 4552 | 10.55 | 1.43 | 1.37-1.48 |
| Additional autoimmune disease with<br>preexisting autoimmunity | 1744 | 38.10 | 1418 | 30.94 | 1.23 | 1.15-1.32 |
| Hashimoto thyroiditis | 2089 | 4.41 | 1474 | 3.11 | 1.42 | 1.33-1.52 |
| Graves' disease | 1696 | 3.52 | 1207 | 2.51 | 1.41 | 1.31-1.51 |
| Psoriasis | 1519 | 3.17 | 1299 | 2.71 | 1.17 | 1.09-1.26 |
| Psoriasis with medication | 488 | 1.00 | 405 | 0.83 | 1.21 | 1.06-1.38 |
| Rheumatoid arthritis | 1175 | 2.43 | 826 | 1.71 | 1.42 | 1.30-1.56 |
| Rheumatoid arthritis with medication | 611 | 1.26 | 421 | 0.87 | 1.45 | 1.28-1.64 |
| Sjögren syndrome | 604 | 1.24 | 419 | 0.86 | 1.44 | 1.27-1.63 |
| Sjögren syndrome with medication | 110 | 0.22 | 72 | 0.15 | 1.53 | 1.14-2.06 |
| Diabetes Type 1 with Insulin | 381 | 0.78 | 304 | 0.62 | 1.25 | 1.08-1.46 |
| Ulcerative colitis | 367 | 0.75 | 282 | 0.58 | 1.30 | 1.12-1.52 |
| Ulcerative colitis with medication | 212 | 0.43 | 175 | 0.36 | 1.22 | 1.00-1.49 |
| Crohn's disease | 298 | 0.61 | 235 | 0.48 | 1.27 | 1.07-1.50 |
| Crohn's disease with medication | 155 | 0.32 | 120 | 0.24 | 1.29 | 1.02-1.64 |
| Polymyalgia | 287 | 0.59 | 233 | 0.47 | 1.24 | 1.04-1.47 |
| Polymyalgia with medication | 174 | 0.36 | 154 | 0.31 | 1.13 | 0.91-1.40 |
| Multiple sclerosis | 271 | 0.55 | 226 | 0.46 | 1.20 | 1.01-1.43 |
| Multiple sclerosis with medication | 99 | 0.20 | 96 | 0.20 | 1.04 | 0.78-1.37 |
| Sarcoidosis | 266 | 0.54 | 124 | 0.25 | 2.14 | 1.73-2.65 |
| Ankylosing spondylitis | 263 | 0.54 | 194 | 0.40 | 1.36 | 1.13-1.63 |
| Ankylosing spondylitis with medication | 82 | 0.17 | 64 | 0.13 | 1.29 | 0.93-1.78 |
| Celiac disease | 229 | 0.47 | 148 | 0.30 | 1.55 | 1.26-1.90 |
| Alopecia areata | 198 | 0.40 | 152 | 0.31 | 1.30 | 1.05-1.61 |
| Vitiligo | 132 | 0.27 | 97 | 0.20 | 1.36 | 1.05-1.77 |
| Immune thrombocytopenic purpura | 83 | 0.17 | 53 | 0.11 | 1.56 | 1.10-2.20 |
| Immune thrombocytopenic purpura with medication | 30 | 0.06 | 15 | 0.03 | 1.96 | 1.06-3.62 |
| Cutaneous lupus erythematosus | 81 | 0.17 | 63 | 0.13 | 1.29 | 0.93-1.80 |
| Systemic lupus erythematosus | 63 | 0.13 | 47 | 0.10 | 1.34 | 0.92-1.95 |
| Systemic lupus erythematosus with medication | 41 | 0.08 | 30 | 0.06 | 1.35 | 0.85-2.16 |
| Arteritis temporalis | 53 | 0.11 | 33 | 0.07 | 1.63 | 1.05-2.53 |
| Arteritis temporalis with medication | 46 | 0.09 | 26 | 0.05 | 1.78 | 1.10-2.89 |
| Bullous pemphigoid | 45 | 0.09 | 29 | 0.06 | 1.54 | 0.97-2.46 |
| Myasthenia gravis | 41 | 0.08 | 27 | 0.06 | 1.50 | 0.93-2.44 |
| Wegener's disease | 41 | 0.08 | 16 | 0.03 | 2.51 | 1.42-4.46 |
| Wegener's disease with medication | 28 | 0.06 | 12 | 0.03 | 2.27 | 1.16-4.44 |
| Primary biliary cirrhosis | 40 | 0.08 | 32 | 0.07 | 1.24 | 0.78-1.97 |
| Autoimmune hepatitis | 38 | 0.08 | 35 | 0.07 | 1.10 | 0.69-1.74 |
| Systemic scleroderma | 35 | 0.07 | 50 | 0.10 | 0.70 | 0.45-1.08 |
| Autoimmune hemolytic anemia | 21 | 0.04 | 15 | 0.03 | 1.37 | 0.71-2.65 |
| Behcet's disease | 21 | 0.04 | 9 | 0.02 | 2.42 | 1.10-5.35 |
| Guillain-Barré syndrome | 20 | 0.04 | 9 | 0.02 | 2.14 | 0.99-4.66 |

Incidence rate (IR) in percent to 1000 person-years without a previous diagnosis of the disease. IRR- Incident rate ratio \*\*Weighted events have been rounded, while the estimates used the exact values

From the list of autoimmune diseases, only those who had at least 20 events in the COVID-19 group were further investigated. Thus, 24 of 64 outcomes were excluded from the analysis. The remaining 40 outcomes represented 30 diseases (Table 2). The most common incident autoimmune diseases with an IR above 1 in 1000 person-years in the COVID-19 group were Hashimoto thyroiditis (IR COVID-19 =4.41, IRR=1.42, 95%-CI=1.33-1.52), Graves’ disease (IR COVID-19=3.52, IRR=1.41, 95%-CI=1.31-1.51), psoriasis (IR COVID-19=3.17, IRR=1.17, 95%-CI=1.09-1.26), rheumatoid arthritis (IR COVID-19 =2.43, IRR=1.42, 95%-CI=1.30-1.56) and Sjögren syndrome (IR COVID-19=1.24, IRR=1.44, 95%-CI=1.27-1.63). In addition, the more specific disease definitions with medication led to similar results (Table 2). The highest IRRs, i.e., the largest significant effect estimates, were found for Wegner’s disease (IRR=2.51, 95% CI=1.42-4.46), Behcet’s disease (IRR=2.42, 95% CI=1.10-5.35), sarcoidosis (IRR=2.14, 95% CI=1.73-2.65) and arteritis temporalis (IRR=1.63, 95% CI=1.05-2.53). Wegner’s disease, Behcet’s disease and arteritis temporalis belong to the group of rare autoimmune diseases involving vasculitis, i.e., small vessel inflammatory processes. Among preselected autoimmune diseases, only systemic scleroderma had IRR estimates below one, but the group differences were not statistically significant (Table 2).

### Incidence of diagnosed autoimmune diseases in subgroups

Regarding population subgroups, the IRR for an incident autoimmune disease for persons with COVID-19 compared to those without COVID-19 did not differ significantly across age groups or between men and women. However, the absolute incidence of any first autoimmune disease per 1000 person-years was considerably higher among older than younger persons, e.g., IR=25.04, IRR=1.42, 95%-CI=1.28-1.57 and IR=19.55, IRR=1.34, 95%-CI=1.17-1.53 among persons with documented COVID-19 in age groups 65-79 years and above 80 years of age compared to IR=4.17, IRR=1.63, 95%-CI=1.29-2.06 among persons with documented COVID-19 below 18 years of age. We also observed a higher absolute incidence of any newly diagnosed autoimmune disease among women with COVID-19 (IR =18.02, IRR=1.42, 95%-CI=1.35-1.48) than among men with COVID-19 (IR =11.33, IRR=1.45, 95%-CI=1.35-1.54) (Figure 2).

**Figure 2.**
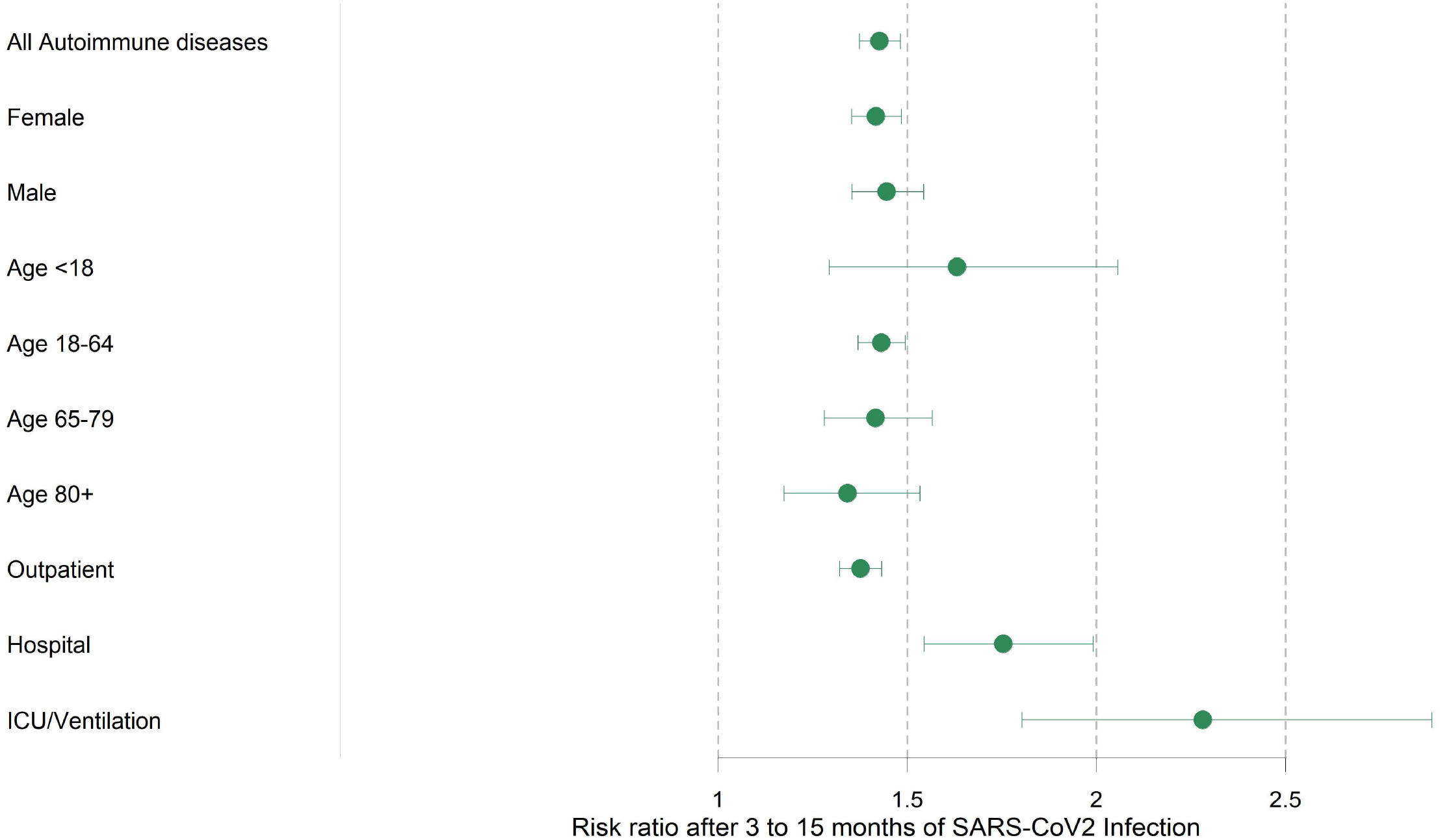
Forest plot comparing incident rate ratios for any first-onset autoimmune disease 3 to 15 months after SARS-CoV-2 infection by subgroup. The severity of COVID-19 was operationalized as only outpatient care, usual hospital care and ICU/ventilation-intensive care unit and mechanical ventilation.

Among individuals with COVID-19, absolute incidence rates as well as IRR relative to controls increased according to the severity of the acute COVID-19 disease, ranging from IR=13.96, IRR=1.38, 95%-CI=1.32-1.43 among persons with outpatient treatment for COVID-19, to IR =28.39, IRR=1.75, 95%-CI=1.54-1.99 among those hospitalized due to COVID-19, and IR =36.96, IRR=2.28, 95%-CI=1.80-2.89 among persons requiring ICU/ventilation (Figure 2).

## Discussion

To our knowledge, this is the largest cohort studies to investigate the association between SARS-CoV-2 infection and the subsequent development of autoimmune diseases. The excess risk for any newly diagnosed autoimmune disease was 4.50 per 1000 person-years in this study. The highest IRRs were found for rather uncommon autoimmune diseases of the vasculitis group. For the more common autoimmune diseases, the highest risks were found for rheumatoid arthritis, Sjögren disease, Graves’ disease and Hashimoto thyroiditis, with an increase of approximately 40% compared to a matched cohort without SARS-CoV-2 infection. Those without a prior autoimmune disease and COVID-19 had a 43% higher likelihood of developing an incident autoimmune disease than controls, while those with any preexisting autoimmune disease and COVID-19 had a 23% higher likelihood of being diagnosed with another autoimmune disease. As expected, absolute incidence rates of any autoimmune disease were higher among women compared to men, among older compared to younger individuals and among those without preexisting autoimmune disease. Comparing persons with and without COVID-19, the IRR increased with the severity of COVID-19 as indicated by hospitalization and particularly by ICU/ventilation treatment versus COVID-19 patients in the outpatient sector. Additionally, a higher IRR for a new-onset autoimmune disease was observed in children and adolescents than in adults with/without COVID-19. However, differences between age groups did not reach statistical significance.

Only other cohorts study was found to investigate the onset of 11 autoimmune diseases after a SARS-CoV2-Infection. It reported a hazard ratio of 1.22 (95%-CI=1.10-1.34). Due to the median follow-up of only 0.29 years only 3 of 11 autoimmune diseases were significant[22].

There are several hypotheses regarding the pathogenesis of post-COVID-19, although different mechanisms most likely underlie the complex clinical picture involving multiple organ systems. Drawing parallels to other post infectious syndromes, possible mechanisms include persistence of the virus or viral or remnants, latent virus reactivation, long-lasting tissue damage due to microclotting and chronic inflammation, and autoimmunity [31]. According to current knowledge, autoimmunity following viral infection may be triggered by mechanisms such as epitope spreading, bystander activation, molecular mimicry, and cryptic epitopes [32]. SARS-CoV-2 shares characteristics of other viruses associated with the development of autoimmunity. Acosta-Ampudia and Anaya summarized these hypotheses as follows. 1. Superantigen activity: The S protein of SARS-CoV-2 contains sequence and structure motifs similar to those of a bacterial superantigen and can bind directly to the T-cell receptor. 2. Molecular mimicry: Accumulating evidence demonstrates that the virus has structural similarity to host-derived components. 3. Neutrophil extracellular trap (NET) formation. 4. Type I interferon (IFN) response. 5. “Overt immunity” which describes the appearance of multiple autoantibodies and diverse autoimmune diseases that are significantly associated with SARS-CoV-2 [33]. These mechanisms are in line with several serological studies demonstrating the onset of IgG autoantibodies[34, 35] or emergence of self-reactive B cells[36] as a response to SARS-CoV-2. Moreover, autoantibodies generated during infection are negatively correlated with SARS-CoV-2 antibodies but positively correlated with hyperinflammation markers during acute illness as well as biomarkers for certain post-acute conditions[37]. These findings highlight a potential link between autoreactivity, severity of COVID-19 and susceptibility to post-acute sequelae. Indeed, serological studies have found persisting patterns of autoreactivity in severe COVID-19 cases even after most autoimmunological markers have subsided after the acute phase [34, 36]. This suggests latent autoimmunity acquired by some patients, which may lead to de novo autoimmune diseases in the long run [23, 35].

Early clinical case studies reported few cases of onset autoimmune diseases following COVID-19. There is growing consensus regarding the relevance of long-term studies on this matter [23, 38]. Two recently conducted systematic reviews and meta-analyses for example show the association between diabetes mellitus and SARS-CoV-2 infection and conclude that the excess risk of type 1 diabetes is small but relevant from a public health perspective although the underlying mechanisms remain to be elucidated in order to prove a causal relationship [39, 40]. Results of the present study regarding the excess risk of newly diagnosed type 1 diabetes in relation to documented SARS-CoV-2 infection are in line with these previous findings.

We also found the overall excess risk for a first autoimmune disease to be 4.50 per 1000 person-years, which is much smaller than previously proposed for other potential chronic sequelae of COVID-19. For cardiovascular diseases, the excess risk was 45.29[13]; for mental diseases, it was 36.48 [14]; and for neurologic disorders, it was estimated to be 70.69 per 1000 people[15]. One reason for this could be that autoimmune diseases are less frequent and the detection time is much longer than that for other diseases. The much larger IRR for hospitalized patients and patients with ICU/ventilation was also reported elsewhere [11, 17].

### Strengths and limitations

The main strength of our analysis is its large dataset including more than 600,000 COVID-19 patients and a minimum follow-up period of three to 15 months. This unselected sample from all over Germany covers both outpatient and inpatient care and thus constitutes a unique and comprehensive source of evidence. Our analysis is based on confirmed diagnoses documented by ambulatory physicians and hospital discharge diagnosis. Accordingly, our results are not subject to possible distortions resulting from selective, incomplete, or inadequate self-reporting of symptoms but instead rely on information provided by medical professionals. To avoid confounding the relationships between outcomes and exposure, we applied matching on relevant covariates, age, sex, previous autoimmune disease and several prevalent diseases and utilization of outpatient and inpatient care. The results were confirmed by the fact that estimates of individual outcome definitions were similar with and without additional consideration of disease-specific medication.

Due to the observational nature of our study, we could not determine a causal interpretation of the results. We could not exclude the possibility that our results were affected by unmeasured confounding, although we minimized differences between the COVID-19 and control cohorts by matching. Vaccination status could not be validly assessed in German claims data. Our results may also have been subject to greater symptom awareness of individuals following SARS-CoV-2 infection or detection bias that may have arisen if the health status of individuals after the onset of COVID-19 was more closely monitored and better documented by physicians. Individuals with a mild or asymptomatic course of COVID-19 were likely to be underrepresented in our study because SARS-CoV-2 infections may not have been documented[41], especially in the first months of the pandemic. The resulting selection of more severe COVID-19 cases may have led to higher incidence estimates in this cohort. In addition, individuals with undocumented SARS-CoV-2 infection may have been included in the control cohort. To the extent that post-COVID also occurred in individuals with undocumented infections, this misclassification induced an overestimation of incidence rates in the control group and, thus, a bias toward the null in estimators of incidence rate ratios.

## Conclusion

In this large matched cohort study, COVID-19 was associated with an increased risk of being newly diagnosed with autoimmune disease 3-15 months after SARS-CoV-2 infection. The strength of the association with SARS-CoV-2 infection was most pronounced for autoimmune diseases in the vasculitis group. A more severe course of COVID-19 was associated with a higher likelihood of being newly diagnosed with autoimmune disease. Incident autoimmune diseases were significantly more common in the post-COVID-19 period in all age and sex groups. The autoimmunity hypothesis is supported by a body of evidence linking viral infections to the pathogenesis of autoimmune diseases as well as results from recent clinical and basic research demonstrating persisting autoantibodies and serological autoreactivity following SARS-CoV-2 infection in a subset of patients. Further epidemiologic, clinical and basic science research is warranted to determine whether SARS-CoV-2 infection triggers the onset of autoimmune disease, to identify the underlying mechanisms and persons at risk, and to investigate effective means of prevention or early treatment.

## Supporting information

RECORD Checklist

## Data Availability

The raw data used in this study cannot be made available in the manuscript, the supplemental files, or in a public repository due to German data protection laws (Bundesdatenschutzgesetz). The aggregated data is stored on a secure drive at ZEGV.

