## Supplementary material for "Incident autoimmune diseases in association with a SARS-CoV-2 infection: A matched cohort study"

**Supplementary material S1 Health outcomes**

| **Name** | **ICD-10 GM Code** | **ATC** |
| --- | --- | --- |
| Addison's disease | E27.1 |  |
| Alopecia areata | L63 |  |
| Autoimmune hemolytic anemia | D59.1 |  |
| Arteriitis temporalis | M31.6 | B01AX, H02AB, H02BX, L01BA, L01XC, L04AA, L04AB, L04AC, L04AD, L04AX, M01CX, P01BA |
| Graves' disease | E05, E06.2 | B01AX, H02AB, H02BX, L01BA, L01XC, L04AA, L04AB, L04AC, L04AD, L04AX, M01CX, P01BA |
| Ankylosing spondylitis | M45.0 | H02AB, H02BX, L01BA, L01XC, L04AA, L04AB, L04AC, L04AD, L04AX, M01CX, P01BA |
| Behcet's disease | M35.2 | B01AX, H02AB, H02BX, L01BA, L01XC, L04AA, L04AB, L04AC, L04AD, L04AX, M01CX, P01BA |
| Churg-Strauss disease | M30.1 |  |
| Morbus Crohn | K50 | A07EA, A07EC, A07EF, H02AB, L01BA, L01BB, L04AA, L04AB, L04AC, L04AD, L04AX, M01CX |
| Dermatomyositis | M33.1 | B01AX, H02AB, H02BX, L01BA, L01XC, L04AA, L04AB, L04AC, L04AD, L04AX, M01CX, P01BA |
| Diabetes type I | E10, | A10AB, A10AC, A10AD, A10AE, A10AF |
| Dermatitis herpetiformis Duhring | L13.0 |  |
| Guillain-Barré-syndrome | G61.0 (only inpatient) |  |
| Goodpasture syndrome | M31.0 | B01AX, H02AB, H02BX, L01BA, L01XC, L04AA, L04AB, L04AC, L04AD, L04AX, M01CX, P01BA |
| Hashimoto's thyroiditis | E06.3 |  |
| Autoimmune Hepatitis | K75.4 |  |
| Juvenile rheumatoid arthritis | M08.1 | H02AB, H02BX, L01BA, L01XC, L04AA, L04AB, L04AC, L04AD, L04AX, M01CX, P01BA |
| Kawasaki syndrome | M30.3 | B01AX, H02AB, H02BX, L01BA, L01XC, L04AA, L04AB, L04AC, L04AD, L04AX, M01CX, P01BA |
| Cutaneous lupus erythematosus | L93 |  |
| Cryoglobulinemia | D89.1 |  |
| Systemic lupus erythematosus | M32 | B01AX, H02AB, H02BX, L01BA, L01XC, L04AA, L04AB, L04AC, L04AD, L04AX, M01CX, P01BA |
| Morphea | L94.0 |  |
| Multiple sclerosis | G35, G36, G37 | H02AB, H02BX, L01DB, L03AB, L03AX, L04AA, L04AC, L04AX, N07XX |
| Myasthenia gravis | G70.0 |  |
| Necrotizing vasculopathy | M31.9 | B01AX, H02AB, H02BX, L01BA, L01XC, L04AA, L04AB, L04AC, L04AD, L04AX, M01CX, P01BA |
| Bullous pemphigoid | L12 |  |
| Pemphigus vulgaris | L10.0,L10.2,L10.2,  L10.3,L10.4 |  |
| Polyarteritis nodosa | M30.0, M30.2, M30.8 | B01AX, H02AB, H02BX, L01BA, L01XC, L04AA, L04AB, L04AC, L04AD, L04AX, M01CX, P01BA |
| Polymyalgia rheumatica | M35.3 | H02AB, H02BX, L01BA, L01XC, L04AA, L04AB, L04AC, L04AD, L04AX, M01CX, P01BA |
| Polymyositis | M33.2 | B01AX, H02AB, H02BX, L01BA, L01XC, L04AA, L04AB, L04AC, L04AD, L04AX, M01CX, P01BA |
| Psoriasis | L40 |  |
| Idiopathic thrombocytopenic purpura | D69.3 | B02AA, B02BX, G03XA, H02AB, J06BA, L01AA, L01CA, L01XC, L04AA, L04AD, L04AX, P01BA |
| Rheumatoid arthritis | M05, M06, M12.3 | D05AC, D05AD, D05AX, D05BA, D05BB, D05BX, H02AB, H02BX, L01BA, L01XC, L04AA, L04AB, L04AC, L04AD, L04AX, M01CX, P01BA |
| Sarcoidosis | D86 |  |
| Sjögren's syndrome | M35.0 | B01AX, H02AB, H02BX, L01BA, L01XC, L04AA, L04AB, L04AC, L04AD, L04AX, M01CX, P01BA |
| Systemic scleroderma | M34.0 | B01AX, C02KX, H02AB, H02BX, L01BA, L01XC, L04AA, L04AB, L04AC, L04AD, L04AX, M01CX, P01BA |
| Takayasu arteritis | M31.4 | B01AX, H02AB, H02BX, L01BA, L01XC, L04AA, L04AB, L04AC, L04AD, L04AX, M01CX, P01BA |
| Ulcerative colitis | K51 | A07EA, A07EC, A07EF, H02AB, L01BA, L01BB, L04AA, L04AB, L04AC, L04AD, L04AX, M01CX |
| Vitiligo | L80 |  |
| Wegener's disease | M31.3 | B01AX, H02AB, H02BX, L01BA, L01XC, L04AA, L04AB, L04AC, L04AD, L04AX, M01CX, P01BA |
| Biliary cholangitis | K74.3 |  |
| celiac disease | K90.0 |  |

**Supplementary material S2 Covariates for the propensity score**

| **Covariate** | **ICD-10 GM Code** | **ATC Code** | **18+ years** | **<18 years** |
| --- | --- | --- | --- | --- |
| Arrhythmia or Atrial fibrillation | I48 |  | x |  |
| Asthma with drug therapy | J45 | D11AH05, H02AB,R03A, R03BA, R03BC, R03C, R03DA, R03DB, R03DC, R03DX05, R03DX08, R03DX09, R03DX10 | x | x |
| Bronchopulmonale Dysplasie | P271 |  |  | x |
| Coronary artery disease / heart attack | I20, I21, I22, I23, I24, I25 | B01AC, C01DA, C01DX11  C01DX12, C07, C08, C09XA53, C09XA54  C09A, C09B, C09C, C09D, C10 | x |  |
| Cerebrovascular or Stroke | G45, G46, H34, I60  I61, I62, I63, I64  I65, I66, I67, I68  I69 |  | x |  |
| Chronic kidney disease | N18, N19, N032, N033, N034, N035, N036, N037, N1880, I131, I120, N052, N053, N054, N055, N056, N057 |  | x |  |
| COPD | J44, J438, J432 | R03A, R03BB, R03C, R03DX07 | x |  |
| Heart_defects | Q20, Q21, Q22, Q23  Q24, Q25, Q26, Q27  Q28, |  |  | x |
| Dementia | F00, F01, F02, F03, F04, G30, G311,G312, G3182, G308, G310 |  | x |  |
| Depression | F32, F33 | N05AN, N06AA, N06AB  N06AF, N06AG, N06AX | x |  |
| Other Diabetes with insulin | E11,E12,E13,E14 | A10A | x | x |
| Other Diabetes without insulin | E11,E12,E13,E14 |  | x |  |
| Dialysis* |  |  | x | x |
| Down syndrome | Q90 |  | x | x |
| Epilepsie | G40,G41 |  |  | x |
| hematological cancer currently not treated | C81, C82, C83, C84  C85, C86, C88, C90, C91, C92, C93, C94  C95, C96 |  | x | x |
| hematological cancer currently treated | As the above row | L01, German Treatment Code (EBM 17370, 17372, 25320, 25321, 25330, 25331, 25332, 25333) | x | x |
| chronic viral hepatitis | B18 |  | x |  |
| heart failure | I099, I110, I130, I132  I255, I42, I43, I500, I501, I509 | C01A, C03, C07, C09A, C09B, C09C, C09D | x |  |
| HIV | B20, B21, B22, B230, B238, B24, U60, U61  U85, Z21 |  | x |  |
| arterial hypertension with drug therapy | I10,I11,I12,I13,I15 | C02, C03, C07, C08, C09 | x |  |
| Immuno-compromising diseases | D570, D571, D572, D573, D578, D68, D70  D71, D72, D73, D76,  D83, D84, D89, D90, I88, M359 |  | x | x |
| immunosuppressive therapy |  | L04, H02AB, H02B | x | x |
| immune blood disease | D77, D80, D81, D82 |  | x | x |
| Interstitial lung disease | E84, J84 |  | x |  |
| mental retardation | F70, F71, F72, F73  F74, F78, F79 |  | x |  |
| metastatic cancer currently not treated | C77, C78, C79 |  | x | x |
| metastatic cancer currently treated | As the above row | L01, German Treatment Code (EBM 17370, 17372, 25320, 25321, 25330, 25331, 25332, 25333) | x | x |
| Obesity | E66 |  | x | x |
| Organ transplant | T860, T861, T862, T863, T864, T865  T868, Z94, Z941, Z942  Z943, Z944, Z946, Z948, Z949 |  | x |  |
| other neurological disease | G10, G11, G13, G14  G20, G210, G21  G22, G23, G24, G25  G26, G31, G320, G328  G36, G370, G40, G70  G71, G80, G81, G82  G83, G9380 |  | x |  |
| psychiatric disorder | F20, F25, F30, F31 | N02CX, N03AF, N03AG  N03AX, N05AA, N05AB, N05AC, N05AD, N05AE  N05AF, N05AG, N05AH  N05AL, N05AN, N05AX | x |  |
| motoric disorder | F82 |  |  | x |
| Severe lung disease | J430, J431, J60  J61, J620, J628, J63, J64, J65, J66, J67, J68, J691, J698, J70 |  | x |  |
| chronic liver disease / cirrhosis | K703, K704, K709, K72, K74 |  | x |  |
| solid cancer currently not treated | C00, C01, C02, C03, C04, C05, C06, C07  C08, C09, C10, C11,  C12, C13, C14, C15, C16, C17, C18, C19  C20, C21, C22, C23, C24, C25, C26, C30, C31, C32, C33, C34, C37, C38, C39, C40, C41, C43, C45, C46, C47, C48, C49, C50, C51, C52, C53, C54, C55, C56, C57, C58, C60, C61, C62, C63, C64, C65, C66, C67, C68, C69, C70, C71, C72, C73, C74, C75 |  | x | x |
| solid cancer currently treated | As the above row | L01, German Treatment Code (EBM 17370, 17372, 25320, 25321, 25330, 25331, 25332, 25333) | x | x |
| Quarters with outpatient physician contact |  |  | x | x |
| Hospital contact |  |  | x | x |

Autoimmune diseases as defined under S1, age and sex have been matched exact. *Dialysis was defined as billed services.
